# Vaccination history for diphtheria and tetanus is associated with less severe COVID-19

**DOI:** 10.1101/2021.06.09.21257809

**Authors:** Jennifer Monereo-Sánchez, Jurjen J. Luykx, Justo Pinzón-Espinosa, Geneviève Richard, Ehsan Motazedi, Lars T. Westlye, Ole A. Andreassen, Dennis van der Meer

## Abstract

**Background:** COVID-19 is characterized by strikingly large, mostly unexplained, interindividual variation in symptom severity. While some individuals remain nearly asymptomatic, others suffer from severe respiratory failure. It has been hypothesized that previous vaccinations for other pathogens, in particular tetanus, may provide protection against severe COVID-19.

**Methods:** We made use of data on COVID-19 testing from 103,049 participants of the UK Biobank (mean age 71.5 years, 54.2% female), coupled to immunization records of the last ten years. Using logistic regression, covarying for age, sex, respiratory disease diagnosis, and socioeconomic status, we tested whether individuals vaccinated for tetanus, diphtheria or pertussis, differed from individuals that had only received other vaccinations on 1) undergoing a COVID-19 test, 2) the outcome of this test, and 3) whether they developed severe COVID-19.

**Results:** We found that individuals with registered diphtheria or tetanus vaccinations were less likely to develop severe COVID-19 than people who had only received other vaccinations (diphtheria OR=0.46, p=3.6×10^−4^; tetanus OR=0.50, p=5.8×10^−4^).

**Discussion:** These results indicate that a history of diphtheria or tetanus vaccinations is associated with less severe manifestations of COVID-19. These vaccinations may protect against severe COVID-19 symptoms by stimulating the immune system. We note the correlational nature of these results, yet the possibility that these vaccinations may influence the severity of COVID-19 warrants follow-up investigations.

---

The coronavirus disease 19 (COVID-19) pandemic is a global health crisis of unprecedented proportions, having caused millions of deaths,^1^ as well as severely disrupting everyday life and crippling the global economy.^2^ While large scale vaccination campaigns are now underway that promise to reduce its chokehold on our societies, it is likely that the underlying virus (SARS-CoV-2), in its current or mutated forms, will remain to significantly influence our lives for the foreseeable future. It is therefore essential that we identify factors that moderate its pathogenic effects.

A particularly striking characteristic of the COVID-19 clinical picture is the considerable interindividual variation. Although estimates vary widely, it is evident that a large proportion of individuals infected with SARS-CoV-2 remain asymptomatic or show only mild flu-like symptoms,^3^ while others develop severe respiratory difficulties that may lead to death.^4,5^ Old age and pre-existing health conditions are strong predictors of poor outcomes, as are certain ethnic backgrounds and male sex,^6,7^ yet a sizeable portion of apparently healthy individuals also suffer severe outcomes. The cause of this variation remains largely unknown, although differences in immune system function are a likely suspect.

One hypothesis, put forth by several researchers independently, is that previous vaccinations for other pathogens may provide protection against severe COVID-19. The tuberculosis vaccine Bacillus Calmette-Guérin (BCG) has been studied in this context, as it is known to generally boost the innate immune response and reduce respiratory tract infections,^8^ with some studies finding a relation between BCG national vaccination programs and COVID-19 mortality rates.^9^ Several other vaccines may ready the immune response with partial specificity, and thereby produce similar protective effects.^10–12^ Indeed, there appear to be protein sequence similarities of proteins in pathogens targeted by common vaccines and SARS-CoV-2,^13,14^ with combination vaccines for the infectious diseases diphtheria, tetanus, and pertussis (DTP vaccine) containing cross-reactive epitopes with SARS-CoV-2.^15^ Additionally, a team of computer scientists applied an artificial intelligence algorithm in an exploratory fashion to millions of biomedical publications, curated assay databases, and protein databanks. Based on the uncovered associations, they generated a hypothesis that tetanus vaccinations may be linked to COVID-19 severity.^16^ More directly, one recent study found a high correlation between SARS-CoV-2 molecular response and DTP vaccine proteins. In this same study, severe disease outcomes were reduced among DTP vaccinated COVID-19 patients.^17^ Together, these clues, coming independently from researchers with distinct backgrounds and with different methodologies, provide an intriguing possibility that DTP vaccinations may explain part of the clinical variability of COVID-19 by modulating the immune response.

In most countries, including the United Kingdom (UK), citizens are now vaccinated against tetanus early in life, often together with diphtheria and pertussis or polio.^18^ Further, five doses of tetanus and diphtheria booster shots later in life are part of the UK immunization program, and shots are also recommended after incurring wounds or when traveling abroad.^19,20^ There is, however, considerable differences in coverage between countries and population groups; in the UK, the majority of adult individuals do not carry protective antibodies for tetanus nor diphtheria, particularly the elderly.^19,21^ It has also been noted that general population vaccination coverages correlate negatively with COVID-19 mortality rates.^12,16^ While claims of causality should be avoided based on such data, cross-reactive immunity could contribute to the greater protection seen for younger individuals, as these have often been recently vaccinated.

Here, we set out to test the hypothesis that DTP vaccinations explain interindividual variation in COVID-19 test results. For this, we used vaccination records of individuals from the large population cohort UK Biobank (UKB), combined with data on COVID-19 testing and related hospitalization, contrasting people vaccinated for DTP versus those vaccinated for other pathogens.

## Methods

### Participants

We used the data from participants of the UKB population cohort, obtained from the data repository under accession number 55392. The composition, set-up, and data gathering protocols of the UKB have been extensively described elsewhere.^22^ UKB has received ethics approval from the National Health Service National Research Ethics Service (ref 11/NW/0382). For this study, we made use of general practitioner (GP) records that had been linked to only a subset of the UKB participants; given the incomplete linking of these records, i.e. missingness, we selected only participants that had at least one vaccination listed in the GP records in the last ten years, as well as complete information on all covariates, which we refer to as the ‘full sample’. The full sample size was n=103,049, with a mean age of 71.46 years (SD=6.92) in March 2020; 54.19% of the sample was female.

### Immunization records

Descriptions of the UK immunization program and vaccination recommendations can be found at https://www.gov.uk/government/publications/immunisation-schedule-the-green-book-chapter-11. We obtained the GP clinical event records linked to the UKB data, downloaded in March 2021. From this, we extracted vaccination information for each participant, which was used to classify them on whether they had received vaccination for tetanus, diphtheria, pertussis, or any other vaccine (influenza, measles, hepatitis, etc.). We upheld strict classifications, only including entries with unambiguous descriptions of specific vaccine administrations. Please see the Supplementary Information for an overview of the search terms that were used to query these records in order to determine the classifications. We used the provided registration dates for the immunizations to only include vaccinations that took place in the last ten years, as the effectiveness of many vaccinations wane over time, which is why booster shots are often recommended every ten years.^23^ Further, this restricts the analyses to the likely more reliable recent registration records. We additionally identified 96 individuals with a BCG vaccination and excluded these, given the substantial amount of literature on a possible relation with COVID-19 severity.

### COVID-19 test results

The COVID-19 test results were downloaded from the UKB data repository in April 2021. At that time, the data contained information on 157,884 tests on 77,222 unique individuals. Given multiple tests for many of the individuals, we selected first the most severe, and then the last positive test, if available. Of these, 13,664 individuals had at least one entry in the vaccination records and complete covariate data.

In accordance with UKB-issued guidance on the analysis of COVID-19 data, we used the inpatient status as indicated in the ‘origin’ field as a proxy for severity. Inpatient status was assigned if the specimen was taken by an emergency care provider, from an inpatient location, or resulting from a hospital-acquired infection. We further assigned any individual that had COVID-19 registered as cause of death (code U07) to the group of positive-tested severe cases.

### Covariates

Differences in registered vaccination, as well as access to health care and lifestyle (e.g., travel and related exposure to pathogens), may relate to socio-economic status (SES), which in turn has been coupled to COVID-19 risk and outcome.^24,25^ We therefore included the Townsend deprivation index as a covariate in our analyses, as previously described.^26^ We flipped the sign of this measure so that higher scores can be interpreted as higher SES.

We additionally covaried for the presence of respiratory disease diagnoses based on the International Classification of Diseases v10, codes starting with a ‘J’ (yes/no, n=21934/81115), to lower the potential influence of pre-existing differences in respiratory difficulties between the vaccine groups.

### Statistical analysis

We made use of a hierarchical set of analyses, whereby we first compared individuals with versus without a specific vaccination on whether they had been tested for COVID-19 (yes/no). Then, among the group of people that had been tested, we compared them on test results (positive/negative). Finally, among those tested positive, we compared whether they differed in proportion of severe cases (severe/not severe). We additionally ran analyses comparing the test results and proportion of severe cases across the full study sample i.e., not hierarchical, providing complementary information on prevalence.

For each of the three sets of analyses we compared individuals that had been vaccinated for tetanus, diphtheria or pertussis in the last ten years to those that did not receive that specific vaccine or any combination with that vaccine included. For instance, we compared participants that received a tetanus vaccine (either by itself or in combination with other vaccines) to all participants that had not received a tetanus vaccine. For an overview of the number of individuals that had received a vaccination, per outcome measure, see Table 1. Vaccinations are very often combined; we were therefore unable to make exclusive groups, i.e. comparing people that only received one of the vaccines to those that had not. For instance, there were only eight people that had received a diphtheria vaccination and not a tetanus vaccination.

**Table 1.** Number of individuals per vaccination group for each of the study outcomes.

|  | Vaccinated |  | Tested |  | Result |  | Severity |  |
| --- | --- | --- | --- | --- | --- | --- | --- | --- |
|  | Yes | No | Yes | No | Positive | Negative | Severe | Not severe |
| Diphtheria | 5,798 | 97,251 | 685 | 5113 | 134 | 551 | 30 | 104 |
| Tetanus | 6,285 | 96,764 | 747 | 5538 | 150 | 597 | 36 | 114 |
| Pertussis | 358 | 102,691 | 27 | 331 | 8 | 19 | 4 | 4 |

All analyses were run in R v4.0. We report the odds ratios as determined through logistic regression on the above described three outcomes, with the independent variables being ‘vaccinated (yes/no)’, ‘age’, ‘sex’, ‘socioeconomic status’, and ‘respiratory disease’.

As we used three predictors (diphtheria, tetanus and pertussis vaccination status) and carried out three main tests (testing, outcome, and severity), we set the Bonferroni-corrected significance level to alpha=.05/9=.006.

We followed up on the main finding from the primary analyses with survival analysis, through Cox proportional hazards models. Here, we used the number of days from the start of the pandemic (set to March 1^st^, 2020) to the date of the COVID-19 test as the time to event, with the event being the registration of a severe COVID-19 case (n=1,185). We censored the registration of a non-severe positive COVID-19 case, assuming that people who overcome COVID-19 with mild symptoms are not at risk to develop severe COVID-19 in the future, and the end of the study period (set to April 1^st^, 2021). We checked for the association of the diphtheria and/or tetanus vaccination status (yes/no) with survival time, covarying for age, sex, and SES. For these analyses we first removed those individuals who died during the study period (n=91), as we did not have the date of death available. Subsequently, we analyzed the survival probability among those participants with a positive COVID-19 test (n=2,692), as well as across all participants (102,958).

Plots were made through the ggplot2 package.^27^ Differences in demographics between the groups were tested through t-tests and chi-square tests, where appropriate. All preprocessing and analysis scripts are available via https://github.com/JenniferMosa/COVID_DTP.

## Results

### Demographics

Among all individuals with a registered vaccination in the last ten years (N=103,049), those tested for COVID-19 (n=13,664; 13.3%) were significantly older (+0.2 years, p=8.8*10^−4^), more often male (+2.8%, p=2.8*10^−9^), and with a lower SES (−0.23, p=1.7*10^−16^) than those not tested. Among those tested, participants testing positive (n=2,692; 19.7%) were younger (−2.5 years, p=2.6*10^−50^) and with lower SES (−0.60, p=2.8*10^−19^) than those testing negative, with no differences in sex distribution (p=.59). Among positive cases, those with severe COVID-19 (n=1,185; 44%) were significantly older (+3.6 years, p=3.7*10^−32^), more often male (11.2%, p=5.8*10^−9^), and had a lower SES (−0.31, p=.01) than those with non-severe COVID-19. We present the demographics split by vaccination group in Supplementary Table 1. These analyses showed that those that received any of the DTP vaccinations in the last ten years were on average younger and had a higher SES than those without these vaccinations.

### Diphtheria

Among individuals tested for COVID-19, those vaccinated for diphtheria tended to be less likely to test positive for COVID-19 than those unvaccinated (OR=0.81), which reached significance when comparing across the full sample (OR=0.75), see Figure 1, on the left. Furthermore, they were significantly less likely to be a severe case (OR=0.46 among those who tested positive, OR=0.45 over the full sample).

**Figure 1.**
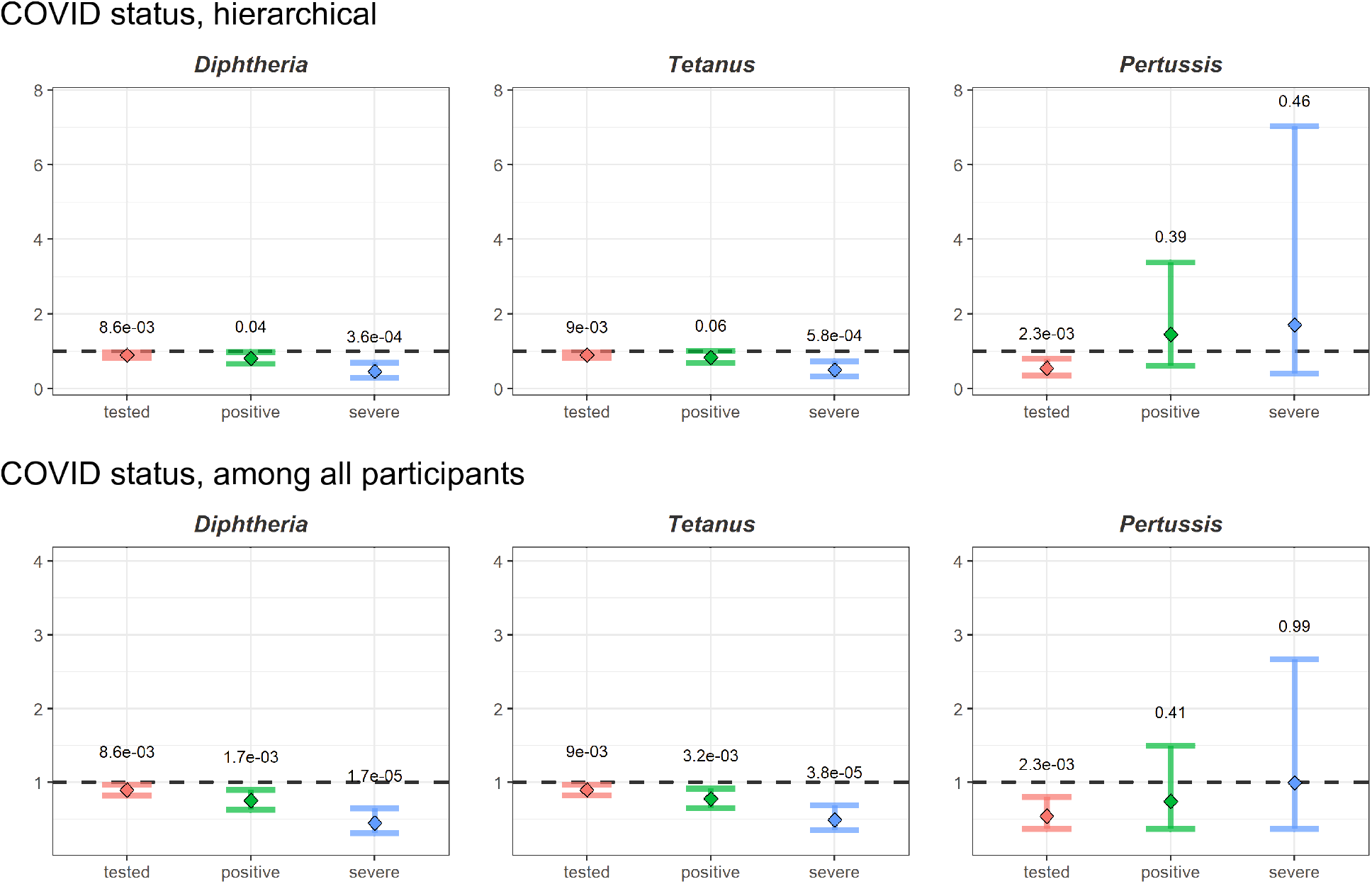
Odds ratios plus 95% confidence intervals (y-axis) for each vaccine group (subplots) comparing whether those that had received a specific vaccine were more or less likely than those not receiving that vaccine to a) being tested for COVID-19, b) getting a positive test result, and c) being a severe case (x-axis and in colors). P-values for each comparison are indicated above the whiskers. The top row presents the hierarchical results, comparing test outcome only in the subset of participants with test data, and case severity only among those testing positive. The bottom row reflects the results over the full sample.

### Tetanus

The findings for tetanus were similar to those for diphtheria, as shown in the middle plots of Figure 1. Individuals receiving this vaccination in the last ten years were less likely to test positive (OR=0.83 among tested, OR=0.78 over full sample). Further, as with diphtheria, those with tetanus vaccinations were half as likely to develop a severe case of COVID-19 (OR=0.50 among positive, OR=0.49 over full sample) than those without this vaccination.

### Pertussis

We found that individuals with pertussis vaccinations were less likely to undergo a COVID-19 test (OR=0.54), while there were no significant differences in the likelihood to test positive or be a severe case, see Figure 1, right side plots. We note the small sample sizes for the analyses of pertussis vaccinations.

### Survival analysis

Given the findings of significant negative associations of diphtheria and tetanus vaccinations with the likelihood of being a severe COVID-19 case, we performed follow-up survival analyses. We drew Kaplan Meier curves for number of days to the development of a severe COVID-19 case, since the start of the pandemic, split by vaccination status for diphtheria and/or tetanus, among those testing positive and across the full samples, as shown in Figure 2. Using Cox regression, we calculated that the survival probability was significantly higher for those with diphtheria and/or tetanus vaccinations than those without, both when testing among COVID-19 cases (hazard ratio=0.62, p=0.01), and across the full sample (hazard ratio=0.45, p=1.4*10^−5^).

**Figure 2.**
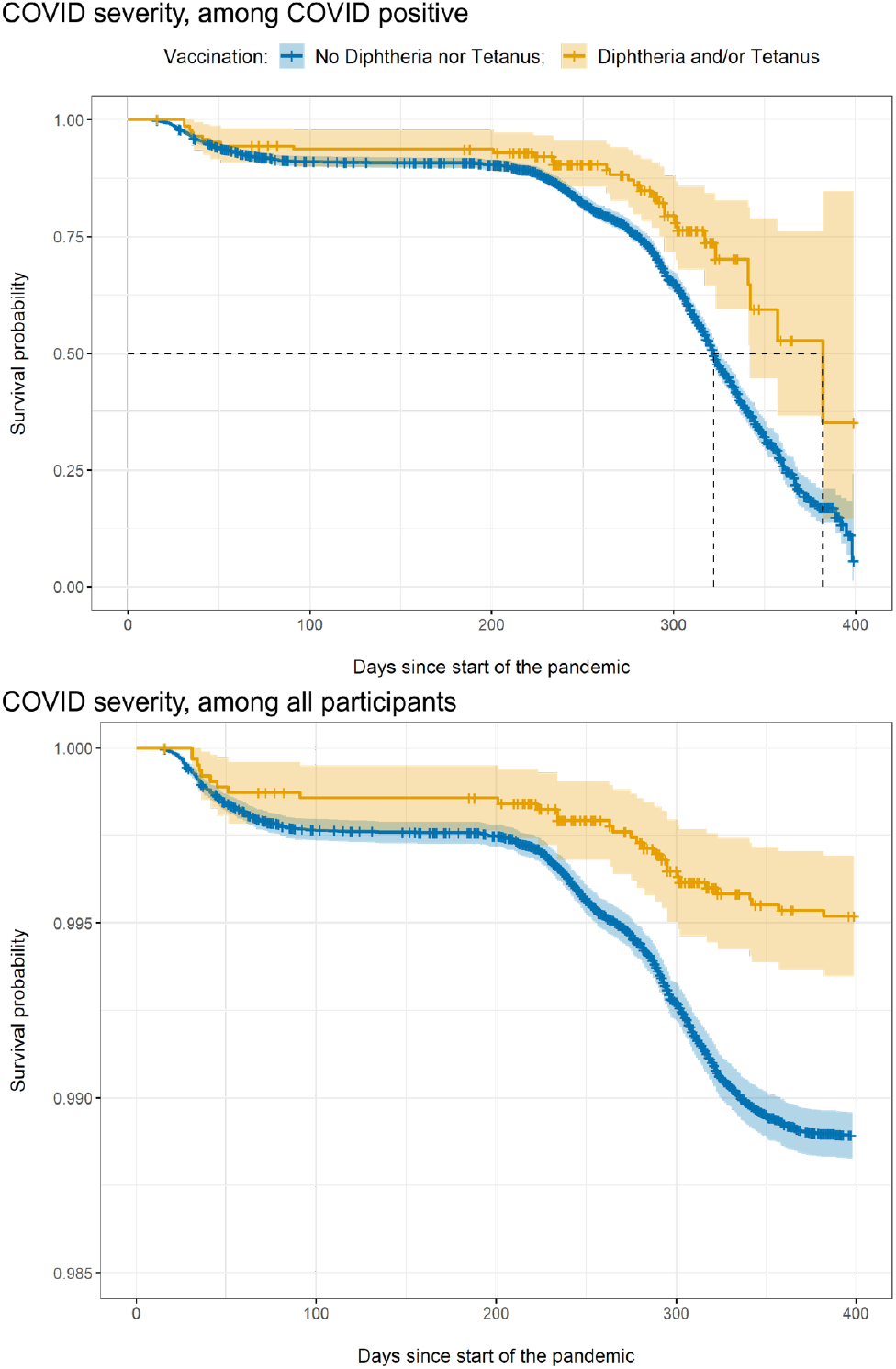
Kaplan Meier survival curves, reflecting the probability over time of developing a severe case of COVID-19 among COVID-19 cases (top) and across the full sample (bottom), split by vaccination status. The x-axis indicates the number of days since the start of the pandemic in the UK, the y-axis indicates the survival probability. The orange line represents the survival probability for those with a diphtheria and/or tetanus vaccination, while the blue line captures the survival probability for those without either of these vaccinations.

## Discussion

Making use of a large-scale elderly population sample, we found that individuals with a diphtheria or tetanus vaccination in the last ten years were half as likely to develop severe COVID-19 than people without these vaccinations. Our follow-up survival analyses further showed that people with diphtheria and/or tetanus vaccinations had a lower risk of developing a severe case, among those tested positive as well as when looking at the full sample.

In line with our *a priori* hypothesis, and replicating recent findings,^17^ our results indicate that DTP vaccinations explain variation in the clinical profile of individuals with exposure to the SARS-CoV-2 virus. Our findings seem to suggest that particularly diphtheria and/or tetanus vaccinations may protect against the development of severe COVID-19 symptoms. One possible mechanism for this would be that these vaccines instill cross-reactive immunity i.e., that they ready the immune response for a SARS-CoV-2 infection, perhaps through protein sequence similarities between the pathogens.^10,15,16^ This is supported by one recent study showing a strong correlation between SARS-CoV-2 response and DTP vaccine proteins.^17^ The potential protective effects were most evident in the analyses of COVID-19 severity, yet they may also explain more subtle differences in testing and test outcomes between those with and without these vaccinations by suppressing mild symptoms, thereby influencing who shows up for testing.

We emphasize that these study outcomes do not allow for any claims of causality. It is possible that the reported associations are driven by an uninvestigated third factor, that distinguishes those with and without registered DTP vaccinations in the last ten years. This could be e.g., lifestyle, international travel, or access to health care, beyond what was captured by the age, sex, respiratory diseases and socioeconomic status covariates. We further note that the vaccination records employed in this study may be incomplete to an unknown extent i.e., vaccinations may not have been recorded correctly or completely. We therefore chose to only compare individuals that had registration of specific vaccinations in the last ten years, thereby avoiding that people with missing data introduce biases in the analyses and minimizing the likelihood that these factors drive the results.

It is likely that the immunity conferred by vaccines wanes over time, and tetanus and diphtheria booster shots have been recommended every decade or when traveling,^23^ although adherence to these recommendations in Europe is low.^19^ Additionally, routine immunization for tetanus was introduced in the UK only in 1961, meaning the majority of individuals in the UKB have not completed their primary course.^18^ Such differences in early vaccination history, in addition to interindividual variation in immune system responses, may further explain differences in COVID-19 clinical profiles. It would be particularly valuable for research investigating the potential mechanistic link between vaccinations and COVID-19 response to use more direct, continuous measures of current immune status than the proxy measure of vaccination history employed here.

We were unable to identify whether either the diphtheria or tetanus vaccination drive the results, as too few individuals received exclusively one of the two vaccines. Further, while we excluded individuals with the BCG vaccine based on its suspected relation with COVID-19 severity, some additional vaccines that were contrasted to the DTP vaccines, e.g. for Measles or Hepatitis B, ^12,14,17^ can potentially have protective effects on COVID-19 symptom presentation as well, thereby obscuring even greater effects of tetanus and/or diphtheria vaccination. It will therefore be important to explore the potential effects of individual vaccines in greater detail, and with more controlled comparison groups.

To conclude, our findings add to literature that specific common vaccines, widely administered to the general population, are associated with severity of COVID-19. These results provide suggestive evidence in favor of what has been previously hypothesized.^10,15,16^ We do not suggest in any way that these vaccines can be seen as substitutes for vaccines developed for COVID-19. However, our findings may still have significant practical implications, by indicating that DTP vaccinations could explain some of the interindividual variation in the response to COVID-19. Based on these promising results, we call for further research into these associations, ideally making use of direct measures of immune activation and well-matched control groups.

## Data Availability

The data incorporated in this work were gathered from public resources. Correspondence and requests for materials should be addressed to.

## Author contributions

J.M.S. and D.v.d.M. conceived the study; J.M.S. and D.v.d.M. pre-processed the data. J.M.S. performed all analyses, with conceptual input from D.v.d.M and E.M.; All authors contributed to interpretation of results; J.M.S. and D.v.d.M. drafted the manuscript and all authors contributed to and approved the final manuscript.

## Acknowledgements

The authors were funded by the Research Council of Norway (276082, 213837, 223273, 204966/F20, 229129, 249795/F20, 225989, 248778, 249795, 298646, 300767), the South-Eastern Norway Regional Health Authority (2013-123, 2014-097, 2015-073, 2016-064, 2017-004, 2019-101), Stiftelsen Kristian Gerhard Jebsen (SKGJ-Med-008), The European Research Council (ERC) under the European Union’s Horizon 2020 research and innovation programme (ERC Starting Grant, Grant agreement No. 802998), ERA-Net Cofund through the ERA PerMed project ‘IMPLEMENT’, and National Institutes of Health (R01MH100351, R01GM104400).

## Supplementary Information

We queried the general practitioner records to identify which individuals had undergone any vaccinations in the last ten years, and specifically whether they had received a tetanus, diphtheria or pertussis vaccination. For this, we performed a search for any matches of the strings ‘vacc’ or ‘immuni’ in these records, followed by searches for matches with the vaccine names or common abbreviations, and then manually checking the resulting lists of matched descriptions. From these, we excluded any description containing keywords such as ‘declined’, ‘did not attend’, ‘no consent’, etc. The generated lists of included and excluded descriptions that capture a vaccination event, per vaccine group, was then triple-checked by two authors (J.M.S. and D.v.d.M.). These lists are presented in the Supplementary Data.

**Supplementary Table 1.** Demographics of the sample, split by whether individuals have received a vaccine (yes/no). Standard deviation is indicated behind the point estimates for age and SES. The test statistic is a t-value for age and SES and a chi-squared value for sex. Behind the test statistic we indicate the p-value behind brackets. SES=socioeconomic status.

| Vaccine |  | Age | Sex | SES |
| --- | --- | --- | --- | --- |
| Tetanus | Yes | 68.22 ± 7.74 | 56.9% | 1.76 ± 2.79 |
|  | No | 71.67 ± 6.81 | 54.0% | 1.37 ± 3.04 |
|  | Test (p) | 34.39 (1.9e-239) | 20.39 (6.3e-06) | -10.70 (1.7e-26) |
| Diphtheria | Yes | 68.23 ± 7.72 | 57.1% | 1.80 ± 2.76 |
|  | No | 71.65 ± 6.82 | 54.0% | 1.36 ± 3.04 |
|  | T (p) | 32.93 (1.1e-219) | 21.61 (3.3e-06) | -11.64 (5.1e-31) |
| Pertussis | Yes | 68.69 ± 7.48 | 56.7% | 1.54 ± 3.00 |
|  | No | 71.47 ± 6.91 | 54.2% | 1.39 ± 3.03 |
|  | T (p) | 7.00 (1.2e-11) | 0.91 (3.4e-01) | -0.96 (3.4e-01) |

